# Long-term serum spike protein persistence but no correlation with post-COVID syndrome

**DOI:** 10.1101/2024.11.11.24317084

**Authors:** Annick Fehrer, Franziska Sotzny, Friederike Hoheisel, Elisa Stein, Laura Kim, Claudia Kedor, Helma Freitag, Cornelia Heindrich, Sandra Bauer, Rebekka Rust, Martina Seifert, Patricia Grabowski, Nina Babel, Carmen Scheibenbogen, Kirsten Wittke

## Abstract

According to the World Health Organization (WHO) and the Centers for Diseases Control and Prevention (CDC), currently an estimated 3 – 6 % of people suffer from post-COVID condition or syndrome (PCS). A subset meets diagnostic criteria for myalgic encephalomyelitis/chronic fatigue syndrome (ME/CFS). Several studies have reported persistence of SARS-CoV-2 proteins or RNA in serum or tissues of both recovered individuals and PCS patients.

In this exploratory study, we investigated whether serum spike protein is associated with PCS and whether it correlates with symptom severity and laboratory biomarkers. We analyzed serum spike protein levels in 121 PCS patients following mild-to-moderate COVID-19, 72 of whom met diagnostic criteria for ME/CFS (post-COVID ME/CFS, pcMECFS). Pre-pandemic seronegative healthy controls (ppHC, n = 32) and post-COVID recovered healthy controls (pcHC, n = 37) after SARS-CoV-2 infection were also included in the study.

We found persistent serum SARS-CoV-2 spike protein in a subset of pcHC (11 %), PCS non-ME/CFS patients (2 %), and pcMECFS patients (14 %). There was no significant association with disease severity, symptoms, or laboratory markers. The spike protein concentration was independent of the time since last spike exposure (infection or vaccination). In five spike-positive out of a total of 22 patients who underwent immunoglobulin depletion via immunoadsorption (IA), spike protein was reduced or completely removed after treatment, indicating binding to immunoglobulins.

In summary, our study identified serum spike protein in a subset of patients after SARS-CoV-2 infection without evidence for a role in the pathogenesis of PCS.

## 1 Introduction

Post-COVID syndrome (PCS) refers to persistent symptoms that occur within three months of a severe acute respiratory syndrome coronavirus type 2 (SARS-CoV-2) infection and last for more than two months according to the World Health Organization (WHO) definition (*1*). It affects individuals across a broad demographic spectrum but is particularly prevalent among young and middle-aged women. PCS is a major burden for health care, economics, and society with an estimated 3 - 6% of patients suffering from chronic and often debilitating illness and no causal therapy so far (*1–4*).

PCS is multisystemic, affects various bodily functions, and presents with a variety of symptoms that include fatigue, exercise intolerance with post-exertional malaise (PEM), cognitive impairments, headaches, muscle pain, and autonomic dysfunction among others. The clinical trajectory can be persistent, relapsing, or fluctuating, significantly impairing the quality of life and work capacity for many affected individuals. A subset of PCS patients meets the Canadian consensus (CCC) or Institute of Medicine (IOM) criteria for the diagnosis of myalgic encephalomyelitis/chronic fatigue syndrome (ME/CFS) (*5–7*).

Although the exact pathomechanisms remain unclear, there are numerous studies providing evidence for inflammation, antibody-mediated autoimmunity, vascular inflammation and circulatory dysfunction, Epstein-Barr-Virus (EBV) reactivation, as well as SARS-CoV-2 persistence in PCS (*8, 9*).

While in the majority of individuals infected with SARS-CoV-2 the virus is successfully cleared from the respiratory tract and blood within one to two weeks in correspondence with resolution of clinical symptoms, a subset of patients experience a prolonged viral persistence or reactivation (*10*). In these patients, recurrent episodes of viral shedding, as well as persisting viral RNA or spike antigen are believed to contribute to continued activation of immune responses, possibly inducing persistent tissue inflammation (*10, 11*). Contrary to common perception of coronavirus disease 2019 (COVID-19) as a respiratory disease, SARS-CoV-2 cell tropism was not only identified in lungs and trachea but also in the kidneys, pancreas, brain, heart, skin, blood vessels, and small intestines (*12*). Apart from primary infection of respiratory epithelium via angiotensin converting enzyme 2 (ACE-2) and co-receptor transmembrane protease serine subtype 2 (TMPRSS2), other cell types like gastrointestinal epithelia, endothelial cells, and neurons may also be susceptible to infection (*10*). SARS-CoV-2 RNA was detected in non-respiratory tissues, including the brain, up to several months after infection (*13, 14*). Furthermore, several studies report a persistent gastrointestinal viral reservoir in some patients with and without persistent symptoms for up to 2 years (*15–17*). Spike protein subunit 1 (S1) was found in CD16+ monocytes of PCS patients up to 15 months after acute COVID-19 (*11*). Peluso et al. showed persistence of SARS-CoV-2 antigen and most commonly spike protein in plasma for more than one year after infection in about 9.2 % of specimens, but longitudinal symptoms were not documented in this study (*18*). Others reported persistent spike protein in the plasma of PCS patients for up to twelve months after infection with a prevalence of up to 60 %, while no spike protein was detected in 26 recovered controls in one study (*19*) or less frequent in recovered controls in another study (*20*). Haddad et al. recently reported an ongoing immune response against SARS-CoV-2 antigens evident from newly activated antibody-secreting cells, suggesting viral persistence or reactivation in 40 % of 61 analyzed PCS patients and none of 25 analyzed recovered controls studied (*21*). However, these studies did not explicitly match time points of SARS-CoV-2 infection or vaccination of their study groups and did not document potential reinfections in the time frame investigated. In addition, studies often lack comprehensive patient characterization and classification as well as correlation with clinical data, or they do not include sufficient controls to contextualize results and verify assay specificity (*17–23*).

To this date, the relationship between PCS and viral persistence is not resolved. Here, we quantified spike protein in serum of PCS patients, as well as pre-pandemic and recovered healthy controls, and analyzed a potential correlation of viral persistence with clinical manifestation and symptom severity. In addition, we investigated the effect of immunoglobulin depletion via immunoadsorption (IA) on serum spike protein in 22 post-COVID ME/CFS (pcMECFS) patients treated in our study as reported elsewhere (*24, 25*).

## 2 Results

### 2.1 Persistent spike protein is detectable in serum of individuals after SARS-CoV-2 infection

SARS-CoV-2 spike receptor-binding domain (RBD) was quantified by ELISA in serum of three post-COVID cohorts: 37 post-COVID recovered healthy controls (pcHC), and 121 PCS patients, of whom 72 met ME/CFS diagnostic criteria (pcMECFS), 1 to 38 months after their last SARS-CoV-2 infection. Additionally, a cohort of 32 pre-pandemic healthy controls (ppHC) was analyzed to validate assay specificity (Fig. 1). While all ppHC serum samples tested negative for SARS-CoV-2 spike protein, 11 % of the pcHC serum samples, 2 % of PCS non-ME/CFS serum samples, and 14 % of the pcMECFS serum samples were positive (Fig. 1A). Both frequency and spike protein concentration (Fig. 1B) did not differ significantly between the three post-COVID cohorts. However, when comparing PCS with pcMECFS patients separately, there was a significant difference in concentration (p=0.0303) and prevalence of spike protein in serum (p=0.0261).

**Fig. 1.**
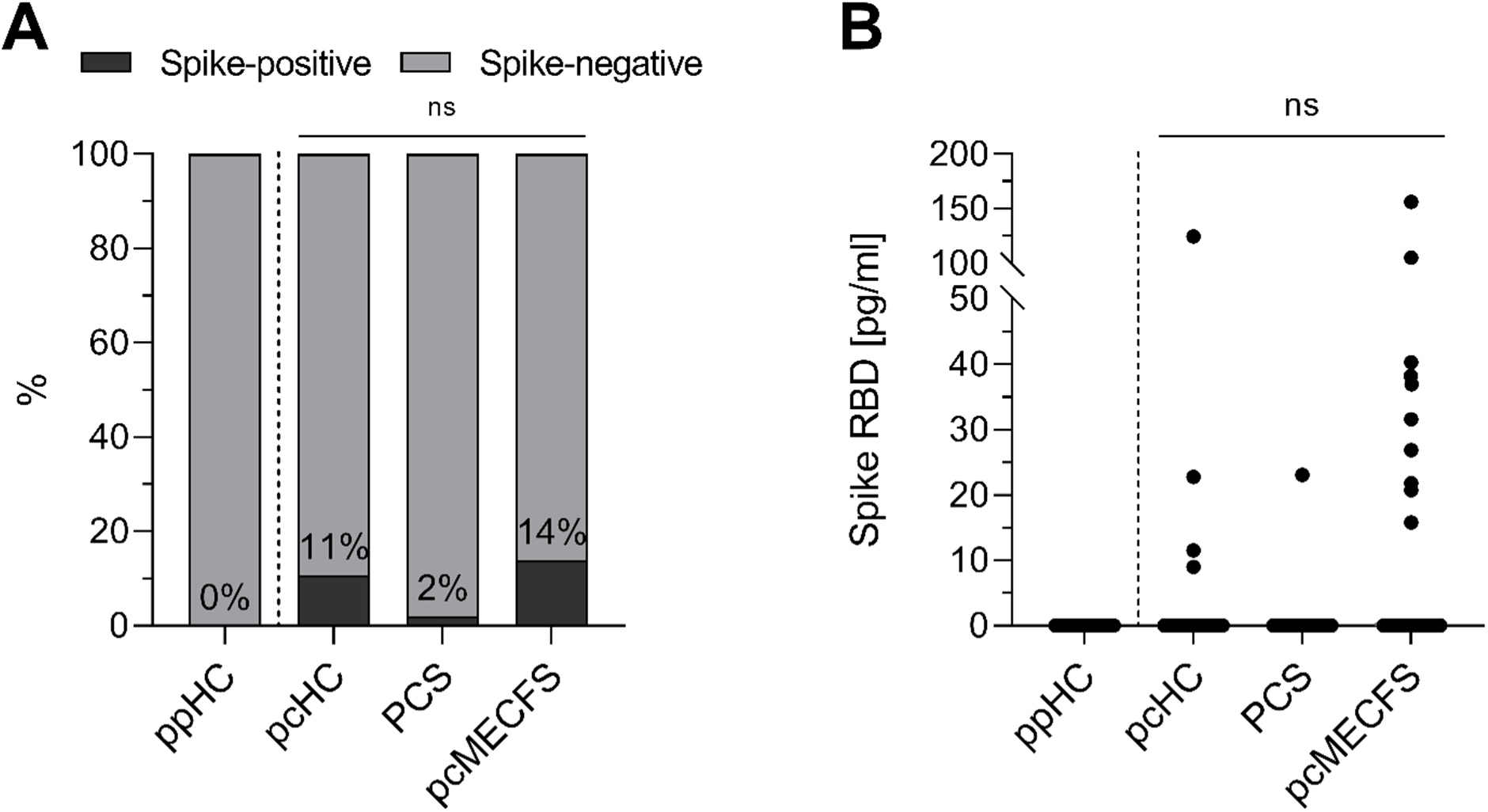
Serum spike RBD concentration of pre-pandemic healthy controls (ppHC), post-COVID healthy controls (pcHC), post-COVID syndrome (PCS) patients, and post-COVID ME/CFS (pcMECFS) patients. Spike RBD was quantified in serum of ppHC (n=32), pcHC (n=37), PCS patients (n=49), and pcMECFS patients (n=72). **(A)** Frequency of spike RBD positive (black) and spike RBD negative (grey) individuals in the different study cohorts. Percentages inside the bars indicate frequency of spike-positive individuals. **(B)** Individual serum spike RBD concentration [pg/ml] of study cohorts. ns=not significant.

### 2.2 Serum spike protein concentration is independent of the time since last SARS-CoV-2 spike protein contact

Next, we examined a potential relationship between SARS-CoV-2 spike protein detection and time since last infection or vaccination in all three post-COVID study groups (pcHC, PCS, pcMECFS). Neither the time since the last SARS-CoV-2 infection (Fig. 2A), time since last SARS-CoV-2 vaccination (Fig. 2B), nor time since last spike protein contact by either infection or vaccination (Fig. 2C) correlated with the serum spike protein concentration. Furthermore, there was no correlation between time since illness-triggering SARS-CoV-2 infection and serum spike protein concentration in PCS and pcMECFS patients (Fig. 2D). Two out of ten spike-positive pcMECFS patients were vaccinated against SARS-CoV-2 shortly before their visit (three and 13 weeks respectively). Among the spike-negative individuals, five pcMECFS and four PCS patients were vaccinated within six weeks before their visit, and four pcMECFS and four PCS patients within 16 weeks before their visit. Two spike-negative patients had a SARS-CoV-2 infection seven or twelve weeks prior.

**Fig. 2.**
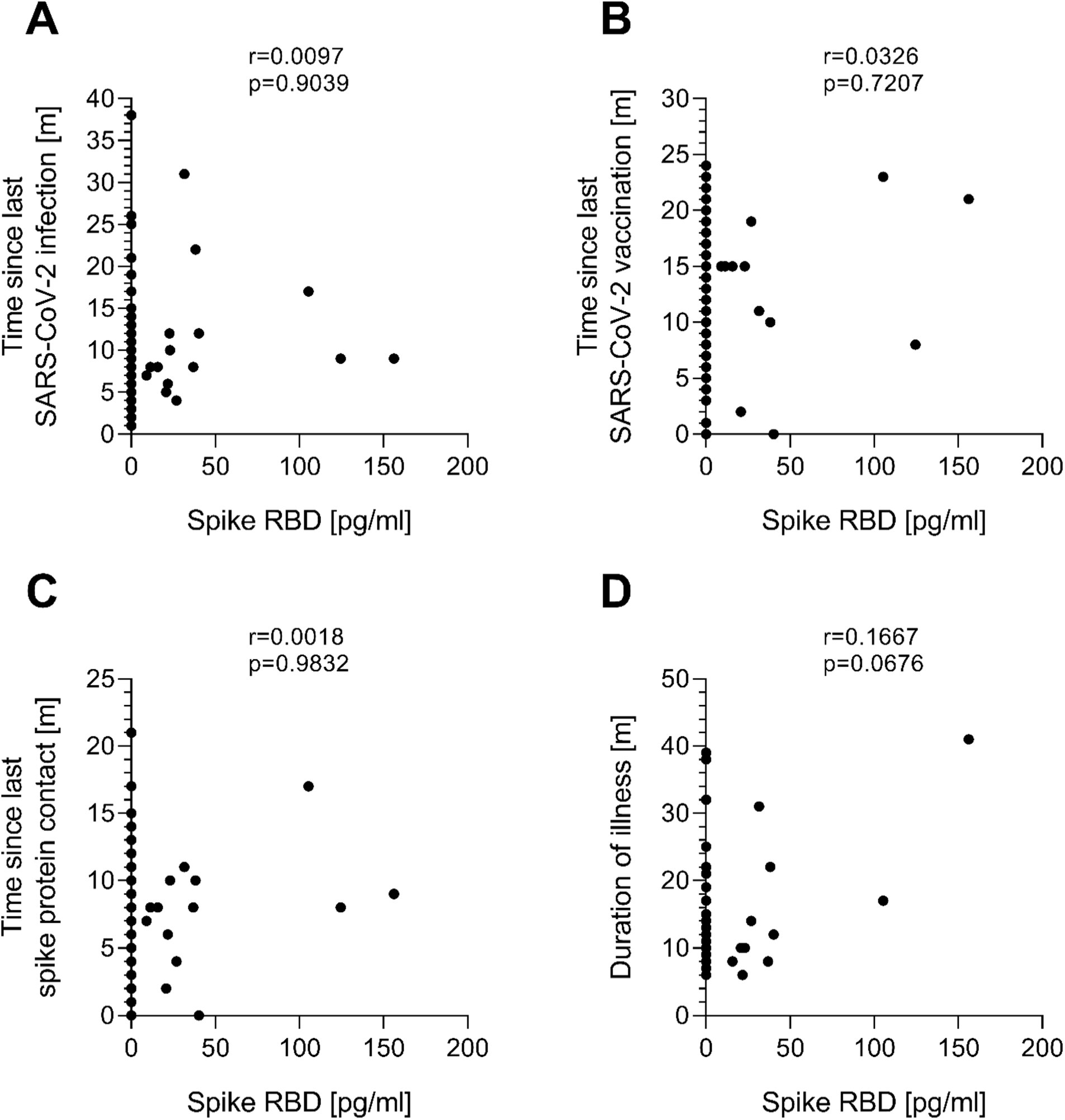
Correlation analysis of spike RBD in serum of post-COVID study groups with time since SARS-CoV-2 infection or vaccination. Spike RBD concentration in serum of pcHC (n=37), PCS patients (n=49), and pcMECFS patients (n=72) was correlated with **(A)** the time since last SARS-CoV-2 infection, **(B)** the time since the last SARS-CoV-2 vaccination, **(C)** the time since the last contact with spike protein either during SARS-CoV-2 infection or vaccination, and **(D)** the duration of illness for patients. Correlation was assessed using Spearman’s rank-order correlation.

### 2.3 Serum spike protein is not associated with pcMECFS disease and symptom severity

Persistent viral reservoirs in PCS is presumed to be associated with inflammation and hypercoagulability, which may contribute to impaired blood flow, endothelial dysfunction, and PCS symptoms (*8, 10, 26*). To evaluate the possible effect of viral persistence in pcMECFS, we investigated a potential association between persisting serum spike protein and severity of disease and key symptoms comparing ten spike-positive and 62 spike-negative pcMECFS patients. We observed no significant difference in severity of disability, assessed by Bell and Short Form Health Survey 36 (SF-36) physical function scores, nor in PEM score, severity of fatigue, pain, cognitive, immune, or autonomic symptoms, as assessed by Composite Autonomic Symptom Score 31 (COMPASS-31), between spike-positive and spike-negative pcMECFS patients (Table 1). The 49 PCS patients were not analyzed as there was only one spike-positive patient in this study group.

**Table 1.** Comparative analysis of disease and symptom severity in pcMECFS patients with (Spike+) or without (Spike-) persistent serum spike RBD.

| Parameter | Spike+ (n=10) | Spike- (n=62) | p-value |
| --- | --- | --- | --- |
|  | median | median |  |
|  | (range) | (range) |  |
|  | n | n |  |
| Severity of disability and PEM |  |  |  |
| Bell Disability Scale | 30.00 | 30.00 | 0.7380 |
|  | (20.00-40.00) | (10.00-60.00) |  |
|  | n=10 | n=61 |  |
| SF-36 Physical Functioning | 25.00 | 35.00 | 0.1021 |
|  | (5.00-50.00) | (0.00-70.00) |  |
|  | n=10 | n=61 |  |
| PEM score | 36.00 | 34.00 | 0.5707 |
|  | (29.00-46.00) | (19.00-46.00) |  |
|  | n=9 | n=58 |  |
| Symptom scores |  |  |  |
| Fatigue score | 8.25 | 8.00 | 0.8686 |
|  | (6.25-10.00) | (5.5-10.00) |  |
|  | n=9 | n=59 |  |
|  | 6.67 | 7.00 |  |
| Cognitive score | (1.67-9.33) | (2.33-10.00) | 0.9265 |
|  | n=9 | n=60 |  |
|  | 2.67 | 3.67 |  |
| Immune score | (1.00-7.33) | (1.00-8.67) | 0.2961 |
|  | n=8 | n=58 |  |
|  | 6.00 | 6.50 |  |
| Headache | (1.00-10.00) | (1.00-10.00) | 0.5623 |
|  | n=9 | n=60 |  |
|  | 6.00 | 7.00 |  |
| Muscle pain | (1.00-10.00) | (1.00-10.00) | 0.6602 |
|  | n=9 | n=60 |  |
|  | 6.00 | 5.00 |  |
| Joint pain | (1.00-9.00) | (1.00-10.00) | 0.8496 |
|  | n=9 | n=60 |  |
| **Assessment of autonomic dysfunction** |  |  |  |
|  | 41.51 | 38.25 |  |
| COMPASS-31 total | (11.55-58.24) | (2.68-76.23) | 0.4045 |
|  | n=10 | n=59 |  |

Mann-Whitney U rank-sum test was performed to test the statistical significance of the difference in numerical data between the study groups. SF-36=Short Form Health Survey 36, PEM=post-exertional malaise, COMPASS-31=Composite Autonomic Symptom Score 31.

A subset of pcMECFS patients had elevated interleukin-8 (IL-8) and a few had elevated D-dimer, platelet count, or mean platelet volume (MPV) as potential markers of endothelial inflammation or hypercoagulation. Levels of these markers were not associated with serum spike persistence in our study cohort (Table 2A). Patients who were treated within the IA study had elevated levels of β2 adrenergic receptor autoantibodies (β2-AdR-AABs). We did not find significant differences in levels of β2-AdR-AABs between spike-positive and spike-negative pcMECFS, nor in levels of anti-S1-IgG (Table 2B). In six of 21 pcMECFS patients we had evidence for endothelial dysfunction assessed by reactive hyperemia index (RHI) using postocclusive reactive hyperaemia peripheral arterial tonometry (RH-PAT), but no association with spike persistence was observed (Table 2C).

**Table 2.** Comparative analysis of routine laboratory and functional markers of pcMECFS patients with (Spike+) or without (Spike-) persistent serum spike RBD.

| Parameter | Reference Range | Spike+ (n=10)<br>median<br>(IQR)<br>n(out of reference<br>range)/n | Spike- (n=62)<br>median<br>(IQR)<br>n(out of reference<br>range)/n | p-value |
| --- | --- | --- | --- | --- |
| **A) Thrombotic and inflammatory markers** |  |  |  |  |
| IL-8 [pg/ml] | <150.00 | 126.80<br>(94.60-132.10)<br>1/7 | 131.00<br>(106.20-176.90)<br>18/55 | 0.2705 |
| D-Dimer [mg/l] | <0.50 | 0.21<br>(0.19-0.34)<br>0/5 | 0.23<br>(0.19-0.38)<br>3/39 | 0.5863 |
| Platelets [/nl] | 150.00-370.00 | 258.50<br>(244.50-277.00)<br>0/8 | 265.50<br>(220.75-308.50)<br>3/60 | 0.9513 |
| MPV [fl] | 7.00-12.00 | 10.90<br>(10.45-11.55)<br>1/8 | 10.70<br>(10.20-11.60)<br>4/60 | 0.5273 |
| **B) β2-AdR-AAB and anti-S1 IgG** |  |  |  |  |
| β2-AdR-AAB [a.u.] | <14.00 | 24.15<br>(10.2-34.79)<br>6/10 | 15.63<br>(8.51-24.50)<br>31/59 | 0.2613 |
|  | 1953.51 | 838.33 | 1185.01 |  |
| anti-S1 IgG [BAU/ml] | (1034.04-3265.13) | (447.05-1957.15) | (547.86-1944.94) | 0.7382 |
| median, (IQR), n | n=16* | n=10 | n=32 |  |

| **C)** | **Markers of endothelial dysfunction** |  |  |  |
| --- | --- | --- | --- | --- |
|  |  | 1.78 | 2.08 |  |
| RHI | >1.67 | (1.41-2.19) | (1.70-2.37) | 0.4531 |
|  |  | 2/4 | 4/17 |  |

\*Median (IQR) of 16 pcHC. Mann-Whitney U rank-sum test was performed to test the statistical significance of the difference in numerical data between the study groups. IQR= interquartile range, NA=not assessed, MPV=mean platelet volume, β2-AdR= β2 adrenergic receptor, AAB=autoantibody, S1=spike protein subunit 1, BAU=Binding Antibody Units, RHI= reactive hyperemia index.

### 2.4 Serum spike protein is reduced by IA and not associated with IA response

Twenty-two of the pcMECFS patients studied here were treated with IA for immunoglobulin depletion (IA-pcMECFS) as reported elsewhere (*24, 25*). Levels of SARS-CoV-2 spike protein were measured before and four weeks after IA (Fig. 3). Interestingly, in all five patients with detectable serum spike protein, levels were either reduced in two or no longer detectable in three patients four weeks after IA. Patient #17 had a SARS-CoV-2 infection immediately before the four weeks after IA visit. Three of the five spike-positive IA patients were responders and two were non-responders as defined by improvement in their physical function (SF-36) four weeks after IA (*24, 25*). Thus, we have no evidence that there is an association between spike protein serum persistence and clinical response to immunoglobulin depletion.

**Fig. 3.**
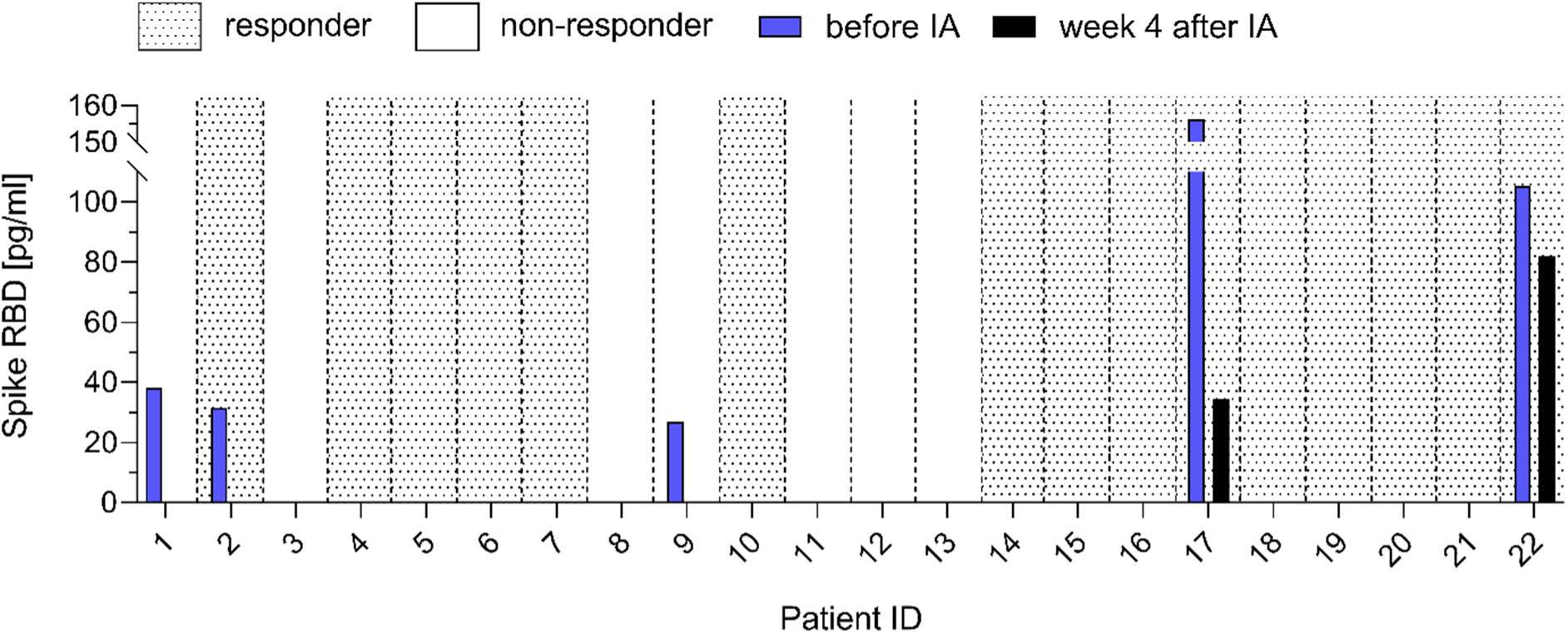
Spike RBD concentration in serum of post-COVID ME/CFS (pcMECFS) patients before immunoadsorption (IA) (IA-pcMECFS) and four weeks after IA. Patients who responded to IA are indicated by dotted black and those who did not by white background based on the improvement in SF-36 physical function four weeks after therapy. Spike RBD concentration [pg/ml] in patients’ serum was determined before (blue bar) and four weeks after IA (black bar).

## 3 Discussion

Our study shows persistence of spike protein in serum up to 31 months after infection in a subset of PCS patients and pcHC. We found neither an association with PCS nor with severity of disease and symptoms in the pcMECFS subgroup. Moreover, we found no link to markers of endothelial dysfunction, inflammation, or hypercoagulation.

The finding from our and other studies of absence of viral proteins in pre-pandemic samples provides strong evidence for the specificity of the detection of viral proteins after SARS-CoV-2 infection in a subset of individuals. The prevalence of antigen persistence in serum in our study is consistent with a previous study by Peluso et al. reporting 9.2 % positivity in 660 pandemic-era plasma samples up to 14 months after SARS-CoV-2 infection (*18*). Other studies report lower or higher levels of viral antigen persistence in serum or plasma ranging from no detectable antigen in both PCS and pcHC study groups to a prevalence of approximately 60 % in PCS patients and 0-30 % in pcHCs (*19, 20, 22, 27, 28*). A recent study reported an association between persistence of SARS-CoV-2 RNA and PCS symptoms up to four months after COVID-19 (*29*). Other studies suggested that tissue viral persistence is associated with PCS symptoms (*17, 30*). Notably, our study shows no association between persistent spike protein and symptom severity in pcMECFS patients 2-38 months after their last SARS-CoV-2 infection. We found spike protein in only 2 % of PCS patients with a significant difference to pcMECFS patients. However, as we detected spike protein in 11 % of pcHC, we consider it more likely to be a random rather than a pathomechanistically relevant finding.

Spike protein can promote microvascular inflammation and thrombogenic processes (*31, 32*), but our study did not find a correlation between endothelial dysfunction, thromboinflammatory markers, and spike protein persistence in pcMECFS patients. In line with our findings, others didn’t observe a link between SARS-CoV-2 persistent shedding in saliva and PCS development or symptoms (*33*). The heterogeneity of findings in the various studies may be related to the generally low levels of viral proteins, potential cross-reactivity with other proteins and different tests that were used.

The hypothesis of viral persistence playing a role in the pathomechanism of PCS is currently used as a rationale for antiviral treatment studies. Paxlovid, a drug consisting of the antiviral agents nirmatrelvir and ritonavir, has been shown to be effective in preventing a severe course of COVID-19 (*34*), which is associated with an increased risk of PCS (*35, 36*). Findings from a study using a large dataset of >281,000 participants from the US Department of Veterans Affairs suggested a protective effect of Paxlovid treatment during acute COVID-19 also for the development of PCS (*37*), while other smaller studies couldn’t reproduce these findings (*38, 39*). A first randomized placebo-controlled trial of 155 PCS patients in the STOP-PASC study showed no significant effect of 15 days of Paxlovid treatment on persistent symptoms (*40*). However, in this study viral particles were not assessed; thus, it is not possible to draw conclusions about the subgroup of patients with evidence of viral persistence.

In addition, our data show a reduction or removal of serum spike protein after immunoglobulin depletion via IA. This suggests that spike protein is bound to anti-spike IgG. However, it cannot be excluded that there is an unspecific binding of spike protein with the antibody-specific sepharose gel matrix used for the IA procedure. The presence or reduction of serum spike protein was not associated with treatment response. This further suggests that persistent spike protein is not causing symptoms in these pcMECFS patients.

The present study has some limitations that need to be addressed. It is a cross-sectional study, that provides a snapshot of serum spike persistence. Temporal profiling of SARS-CoV-2 antigens in the plasma of PCS patients shows fluctuating concentration profiles of both spike and nucleocapsid protein over a time period of up to twelve months after infection (*19*). Regular blood sampling within a defined time frame and interval would be necessary to shed further light on the mechanisms of viral persistence and its temporal detectability in serum.

We determined the concentration of spike protein RBD in the serum. This is worth mentioning, as a previous study reported discrepancies between e.g. S1 and full spike protein detection (*19*). In addition, spike protein may not only be present in a soluble form, but may also be transported in extracellular vesicles, as previously reported (*20, 41*). Other studies have shown persistence of viral RNA in tissues (*13–17*). This should be taken into account for interpretation of the results presented here.

As PCS is a highly heterogeneous disease, viral persistence may play a role only in the pathogenesis of certain subtypes of PCS. Our study focused on a subgroup of patients with ME/CFS or similar symptom complexes. Further research is needed to elucidate potential differences in spike persistence in clinically and immunologically distinct PCS subtypes and its potential relevance for the diagnosis and treatment of patients.

In conclusion, despite the limitations described, our study provides evidence for serum spike persistence in a subset of individuals after SARS-CoV-2 infection without an association with PCS.

## 4 Material and Methods

### 4.1 Study design, study cohorts, and symptom assessment

We conducted an exploratory, cross-sectional study investigating serum spike persistence in individuals with and without PCS symptoms up to 38 months after SARS-CoV-2 infection. The study aimed to determine whether serum spike protein is associated with PCS symptoms or correlates with disease severity and selected laboratory markers.

Serum samples and clinical data from 121 PCS patients suffering from moderate to severe fatigue and exertion intolerance following mostly mild COVID-19 were collected between October 2020 and January 2024 at the ME/CFS outpatient clinic of the Institute of Medical Immunology, Charité, Berlin. From these, 72 met the CCC for diagnosis of ME/CFS (pcMECFS) (*42, 43*). Relevant cardiac, respiratory, neurological, or psychiatric comorbidities were excluded. We analyzed 22 pcMECFS patients who had participated in an observational study of immunoglobulin depletion via IA (IA-pcMECFS) (*24, 25*). Of these, serum samples at timepoints before and four weeks after IA were analyzed.

Additionally, serum samples from 37 pcHC five to twelve months after their last SARS-CoV-2 infection were obtained between January 2023 and October 2023. pcHCs did not suffer from a disease with relevant impairment or take regular medication, and had no infection or vaccination within the last four weeks. To control for assay specificity, 32 pre-pandemic serum samples from ppHC, collected between January 2019 and October 2019, were analyzed.

Cohort characteristics are listed in Table 3. The IA-pcMECFS study group differed significantly from the other study groups due to a longer disease duration, as well as longer time after SARS-CoV-2 infection and vaccination. Severity of disease and symptoms were quantified using questionnaires. PEM was assessed according to Cotler et al. 2018 with scores ranging from 0 to 46 (no to frequent, severe, long lasting PEM) (*44*). Functional ability was scored using the Bell Disability Scale ranging from 0 to 100 (total loss of self-dependence to no restrictions) (*45*). Daily physical activities were assessed using the SF-36 ranging from 0 to 100 (greatest to no restrictions) (*46*). The severity of the cardinal symptoms fatigue, cognitive impairment, immune symptoms, and pain were quantified using a Likert scale (1 = no symptoms to 10 = most severe symptoms). Symptoms of autonomic dysfunction were assessed using the COMPASS-31 ranging from 0 to 100 (no to strongest symptoms) (*47*). As evident from the Bell score, patients with pcMECFS suffered from more severe disease compared to PCS non-ME/CFS patients, while clinical presentation of pcMECFS and IA-pcMECFS study groups were comparable (Table 3). Ethical Committee approval for this project was obtained in accordance with the 1964 Declaration of Helsinki and its later amendments (EA2/066/20 and EA2/067/20). All study participant signed informed consent before study inclusion.

**Table 3.**
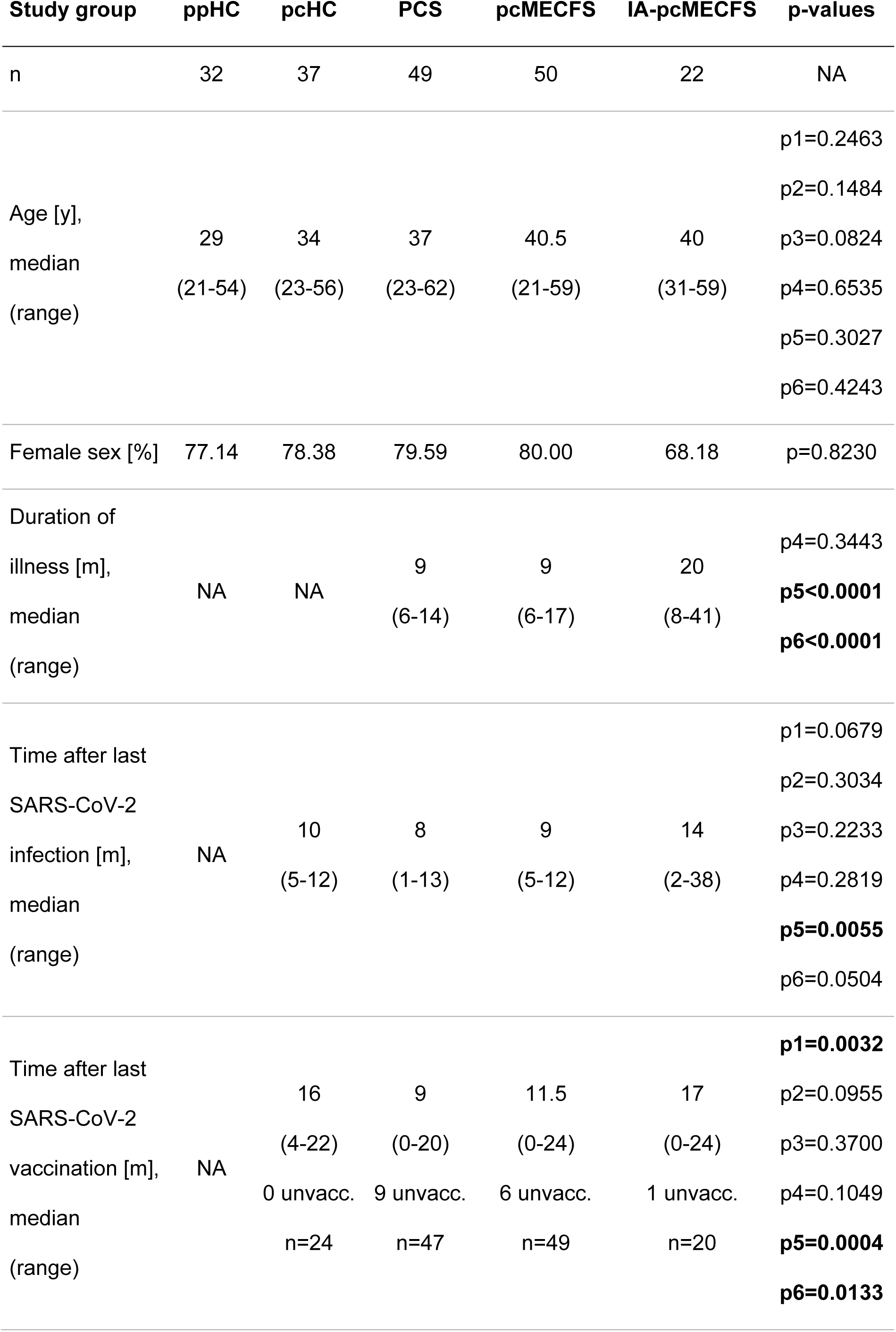
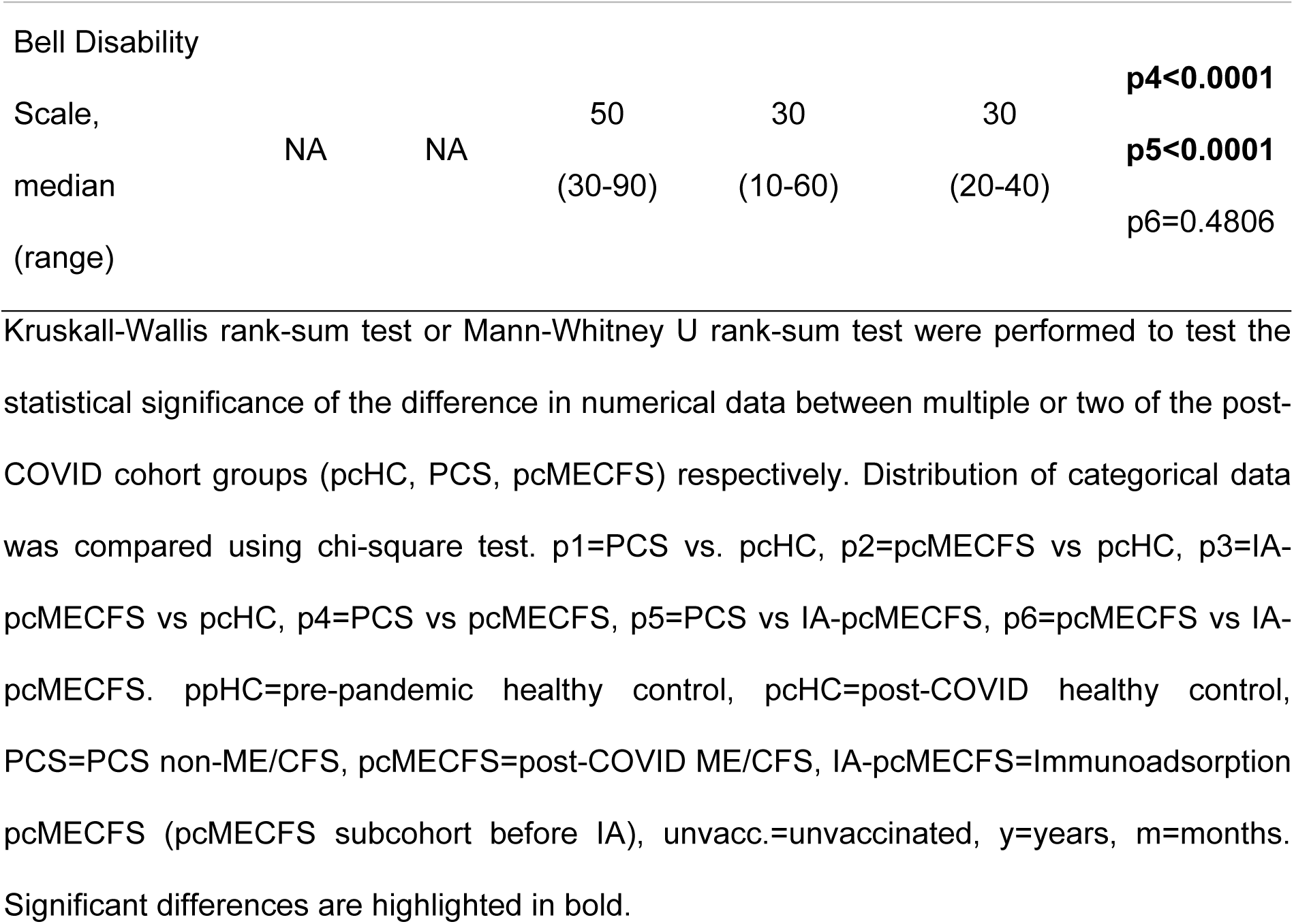
Cohort characteristics

| Study group | ppHC | pcHC | PCS | pcMECFS | IA-pcMECFS | p-values |
| --- | --- | --- | --- | --- | --- | --- |
| n | 32 | 37 | 49 | 50 | 22 | NA |
| Age [y],<br>median<br>(range) | 29<br>(21-54) | 34<br>(23-56) | 37<br>(23-62) | 40.5<br>(21-59) | 40<br>(31-59) | p1=0.2463<br>p2=0.1484<br>p3=0.0824<br>p4=0.6535<br>p5=0.3027<br>p6=0.4243 |
| Female sex [%] | 77.14 | 78.38 | 79.59 | 80.00 | 68.18 | p=0.8230 |
| Duration of<br>illness [m],<br>median<br>(range) | NA | NA | 9<br>(6-14) | 9<br>(6-17) | 20<br>(8-41) | p4=0.3443<br>**p5<0.0001**<br>**p6<0.0001** |
| Time after last<br>SARS-CoV-2<br>infection [m],<br>median<br>(range) | NA | 10<br>(5-12) | 8<br>(1-13) | 9<br>(5-12) | 14<br>(2-38) | p1=0.0679<br>p2=0.3034<br>p3=0.2233<br>p4=0.2819<br>**p5=0.0055**<br>p6=0.0504 |
| Time after last<br>SARS-CoV-2<br>vaccination [m],<br>median<br>(range) | NA | 16<br>(4-22)<br>0 unvacc.<br>n=24 | 9<br>(0-20)<br>9 unvacc.<br>n=47 | 11.5<br>(0-24)<br>6 unvacc.<br>n=49 | 17<br>(0-24)<br>1 unvacc.<br>n=20 | **p1=0.0032**<br>p2=0.0955<br>p3=0.3700<br>p4=0.1049<br>**p5=0.0004**<br>**p6=0.0133** |
| Bell Disability |  |  |  |  |  |  |
| Scale, |  |  | 50 | 30 | 30 | **p4<0.0001** |
| median | NA | NA | (30-90) | (10-60) | (20-40) | **p5<0.0001** |
| (range) |  |  |  |  |  | p6=0.4806 |

Kruskal-Wallis rank-sum test or Mann-Whitney U rank-sum test were performed to test the statistical significance of the difference in numerical data between multiple or two of the post-COVID cohort groups (pcHC, PCS, pcMECFS) respectively. Distribution of categorical data was compared using chi-square test. p1=PCS vs. pcHC, p2=pcMECFS vs pcHC, p3=IA-pcMECFS vs pcHC, p4=PCS vs pcMECFS, p5=PCS vs IA-pcMECFS, p6=pcMECFS vs IA-pcMECFS. ppHC=pre-pandemic healthy control, pcHC=post-COVID healthy control, PCS=PCS non-ME/CFS, pcMECFS=post-COVID ME/CFS, IA-pcMECFS=Immunoadsorption pcMECFS (pcMECFS subcohort before IA), unvacc.=unvaccinated, y=years, m=months. Significant differences are highlighted in bold.

### 4.2 Quantification of serum spike RBD and anti-S1 IgG

Whole blood samples from participants were allowed to clot at room temperature for at least 30 min and then centrifuged at 2000 x g for 10 min at room temperature. Serum was collected and stored at −80°C. Serum concentration of the SARS-CoV-2 spike RBD was determined using Human SARS-CoV-2 RBD ELISA Kit purchased from Thermo Fisher Scientific (EH492RB) and conducted according to manufacturer’s instructions. Serum anti-S1 IgG was quantified using anti-SARS-CoV-2 QuantiVac ELISA (IgG) purchased from Euroimmun (EI 2606-9601-10 G) and conducted according to manufacturer’s instructions. Obtained arbitrary units (AU) were converted into the WHO International Standard (IS) for anti-SARS-CoV-2 immunoglobulin binding activity (Binding Antibody Units, BAU) by multiplying by a factor of 3.2.

### 4.3 Assessment of routine laboratory and functional parameters

D-Dimer, platelet count, MPV, and IL-8 (post erythrocyte lysis) were determined at the Charité diagnostics laboratory Labor Berlin GmbH (Berlin, Germany). Autoantibodies (AABs) against the G protein coupled receptor (GPCR) β2 adrenergic receptor (β2-AdR), were determined by CellTrend GmbH (Luckenwalde, Germany) using indirect ELISA technology.

Peripheral endothelial function was assessed in IA-pcMECFS patients by the RHI using RH-PAT (endoPAT2000, Itamar Medical Ltd., Caesarea, Israel) as previously described (*48*).

### 4.4 Data Collection and Management

Study data, including clinical and routine laboratory parameters, were collected and managed using REDCap electronic data capture tools hosted at Charité—Universitätsmedizin Berlin (*49, 50*).

### 4.5 Statistical analysis

Study data are presented as median with range or interquartile range (IQR) for each study group (n = number of individuals in the study group) as indicated. Nonparametric statistical methods were used. Univariate comparisons of two independent groups were done using the Mann-Whitney-U test and of multiple independent groups using the Kruskal–Wallis test with Benjamini-Hochberg correction of multiple comparisons. Distribution of categorical data was compared using chi-square test. Correlation was assessed using Spearman’s rank-order correlation. A two-tailed p value <0.05 was considered significant. Microsoft Excel 2016 was used for data analysis and GraphPad Prism 9 was used for statistical analysis and graphical presentation.

## Data Availability

All data associated with this study are present in the paper. Correspondence and requests should be addressed to AF.

## Acknowledgements

We would like to thank the patients who participated in this study, as well as Anja Hagemann, Silvia Thiel, and Beate Follendorf for their work in patient care and data management.

## Funding

This study was funded by the ME/CFS Research foundation and the Lost Voices foundation. AF received a scholarship from the Fridericus foundation.

## Author contributions

AF conducted the laboratory work and data analysis of the study. ES, LK, CK, RR, PG, KW, and CS diagnosed and enrolled the patients, and collected the biosamples. AF, FH, and SB collected biosamples of healthy controls and processed all biosamples. FS, HF, and CH managed clinical and routine laboratory data. FS, KW, and CS provided concept, resources, and supervision. AF, FS, and CS interpreted and discussed the results. AF, FS, MS, NB, CS, and KW provided scientific insights. AF, FS, and CS wrote the manuscript. All authors have read, revised, and approved the final version of the manuscript.

## Competing interests

The authors declare no conflict of interest.

## One sentence summary

Our study identified serum spike protein in a subset of patients after SARS-CoV-2 infection without evidence for a role in the pathogenesis of PCS.

## Notes

### Competing Interest Statement

The authors have declared no competing interest.

### Author Declarations

The Ethics Committee of Charite - Universitaetsmedizin Berlin gave ethical approval for this work in accordance with the 1964 Declaration of Helsinki and its later amendments (EA2/066/20 and EA2/067/20).

